# Effects of Specific Protective Resources on the Burnout Levels of Selected Visayan Medical Students from Online-Based Education during COVID-19 Pandemic: A Cross-Sectional Study

**DOI:** 10.1101/2023.08.28.23294675

**Authors:** Jordan Joy Arpilleda, Julia Patricia Bunal, Margaret Therese Rose Montano, Lesly Marie Torrenueva, Ma. Jobelle Acebes, Margaret Angeli Homecillo, Juan Paolo Salvador, Frances Gabrielle Tan, Zackaree Michael Villanueva

**Affiliations:** Cebu Institute of Medicine, Philippines

**Keywords:** Burnout, grit, religiosity, social support, tolerance for ambiguity

## Abstract

**Background:** Burnout is a phenomenon characterized as a consistent state of being exhausted physically, mentally, and emotionally. Grit, tolerance for ambiguity, religiosity, and social support are protective factors that may mitigate burnout and improve life satisfaction. This study assessed the association between specific protective resources of students from a medical school in Visayas at all year levels with online-based education-related burnout during the COVID-19 pandemic.

**Methods:** A total of 234 medical students from a medical school in Visayas during the academic year 2020-2021 were sampled using stratified random sampling technique. Demographics were obtained. Specific protective resources of grit, religiosity, social support and tolerance for ambiguity, and burnout symptoms were measured using validated online questionnaires. Protective associations of specific resources on burnout levels were assessed using multivariate logistic regression analysis. Predictive capabilities of resources with statistically significant protective effects were summarized using Receiving Operating Characteristic (ROC) curve.

**Results:** All year levels experienced burnout based on Maslach Burnout Inventory-Student Survey (MBI-SS) subscales, with PBL 2 having the highest incidence comprising 33 respondents (49%). Majority of the students experienced moderate burnout on emotional exhaustion (44%) and low burnout on depersonalization (58%), while all of them had high burnout levels on the subscale of personal accomplishment (100%). Most students were moderately gritty (91%) and had high tolerance for ambiguity (98%), overall religiosity index (82%), and social support from significant others (68%); family (68%); and friends (76%). Grit, religiosity and social support had positive significant correlations and reductive effects to burnout. Tolerance for ambiguity did not show any significant relationship with burnout. Grit, religiosity and social support are good predictors of burnout. Grit had good diagnostic accuracy and discrimination. Religiosity and social support were moderately accurate predictors of burnout.

**Conclusions:** Grit, religiosity and social support are significantly protective on burnout levels of medical students from online-based education during the COVID-19 pandemic.

## INTRODUCTION

### Background of the Study

The World Health Organization (WHO) included burnout in the 11th Revision of the International Classification of Disease (ICD-11) as a phenomenon characterized as a consistent state of being exhausted physically, mentally, and emotionally.^1^

Factors that may mitigate burnout, affect psychological well-being, and improve life satisfaction include grit defined as the firmness of the mind or spirit and having unyielding courage in the face of hardship or danger.^1^ A meta-analysis by Kim et al also showed that social support from families and peers has a negative correlation with student burnout making it a protective factor.^6^ Religiosity and having a strong spiritual belief and background have protective effects against stress and burnout.^7,8^ Lastly, a low tolerance for ambiguity or uncertainty could be a predictor for burnout as seen in the study done on physicians by Kuhn et al.^9^

Distress during medical school can lead to burnout with significant consequences especially if it continues to occur during residency and further studies.^1^ Moreover, there are still no reports associating the effects of specific protective resources on burnout levels of medical students taking virtual classes during the COVID-19 pandemic. Thus, this study was conducted.

### Statement of the Problem and Objectives

This study primarily aims to assess the association between specific protective resources of students from a medical school in Visayas in all year levels with online-based education-related burnout during the COVID-19 pandemic. Specifically, this study aims to answer the following objectives:

1. Identify the socio-demographic profile of the respondents in terms of: Sex, Age, Year level, Civil/Marital Status, and Religion
2. Determine the level of burnout of the respondents across all year levels in online-based education in terms of: Emotional Exhaustion, Personal accomplishment, and Depersonalization
3. Determine the extent of specific protective resources of all year levels in terms of: Grit, Tolerance for ambiguity (TFA), Religiosity, and Social Support
4. Assess if there is a significant relationship between the specific protective resources and the burnout levels of medical students
5. Assess if grit, tolerance for ambiguity (TFA), religiosity, or social support is predictive of online-based education burnout reduction in medical students
6. Determine the prognostic utility of the specific protective resources in predicting burnout reduction among the medical students

## MATERIALS AND METHODS

The study is a descriptive cross-sectional type. Data collection was done through online questionnaires in Google Forms sent via email to enrolled Problem-Based Learning (PBL) medical students at all year levels in a medical school in the Visayas during the Academic Year 2020-2021. Stratified randomized sampling technique was done. The study formulated survey questionnaires consisting of three parts: (1) demographic characteristics such as age, sex, relationship status, religion, and year level; (2) the validated Maslach Burnout Inventory-Student Survey (MBI-SS), which is a gold standard for defining burnout ^3^; and (3) questionnaires assessing the specific protective resources against burnout including the 5-item Grit scale, Duke University Religion Index (DUREL), Multidimensional Scale of Perceived Social Support (MSPSS), and 16-item Tolerance for Ambiguity Scale (TAS). All hypothesis testing was performed using a 0.05 level of significance. All variables that recorded a significant predictor of burnout reduction were subjected to a Receiver Operating Characteristic (ROC) analysis to plot the sensitivity and 1-specificity of the variables towards diagnosing reduction in burnout. The Area Under Curve was obtained to determine its prognostic utility as a specific protective resource to burnout. Multivariate linear regression analysis was done on grit, religiosity, social support, and TFA to assess predictive associations against burnout. All statistical analyses were performed with GNU PSPP Statistical Analysis Software. Statistical significance was set p < 0.05.

## RESULTS AND DISCUSSION

Among the 234 respondents, eighty-nine percent (89%) are between 21 to 25 years old. Additionally, PBL 2 comprised the bulk of the sample size (29%) while PBL 1 had the least number of respondents (19%) due to the reduction of enrollment rate. Most of the respondents are females (65%), Roman Catholics (82.05%), and single (74%).

Descriptive statistics for survey instruments are shown in Table 1. Burnout, religiosity, and social support are listed by their respective subscales.

**Table 1.** Summary Of Survey Instrument Results (Mean ± Standard Deviation) At 1 Study Sampling Time (N=234).

| Survey Instrument | T1 |
| --- | --- |
| **Burnout** |  |
| Emotional Exhaustion | 22 ± 0.97 |
| Personal Accomplishment | 10.9 ± 0.82 |
| Depersonalization | 5.5 ± 1.01 |
| **Tolerance For Ambiguity** | 5.5 ± 1.02 |
| **Religiosity** |  |
| Organizational Religious Activity (Ora) | 3.8 ± 1.40 |
| Non-Organizational Religious Activity (Nora) | 3.0 ± 1.70 |
| Intrinsic Religiosity (Ir) | 11.0 ± 1.13 |
| **Grit** | 3.2 ± 1.08 |
| **Social Support** |  |
| Significant Other | 5.4 ± 1.80 |
| Family | 5.6 ± 1.00 |
| Friends | 5.7 ± 1.30 |
| Overall | 5.6 ± 1.37 |

T1: 10th May - 31st May 2021

Table 2 depicts the levels of burnout in terms of their online classes in the different subscales of MBI-SS across all year levels A.Y. 2020-2021. Majority of the PBL 1 and PBL3 respondents had moderate burnout levels on the emotional exhaustion subscale and low burnout levels on depersonalization. Most of the PBL 2 respondents had high burnout levels on emotional exhaustion but low burnout levels on depersonalization. Most PBL 4 respondents had low burnout scores on both emotional exhaustion and depersonalization. All respondents had high levels on the personal accomplishment subscale. An increased degree of burnout is present if the total values of emotional exhaustion and depersonalization are high and the value of personal accomplishment is low.

**Table 2.**
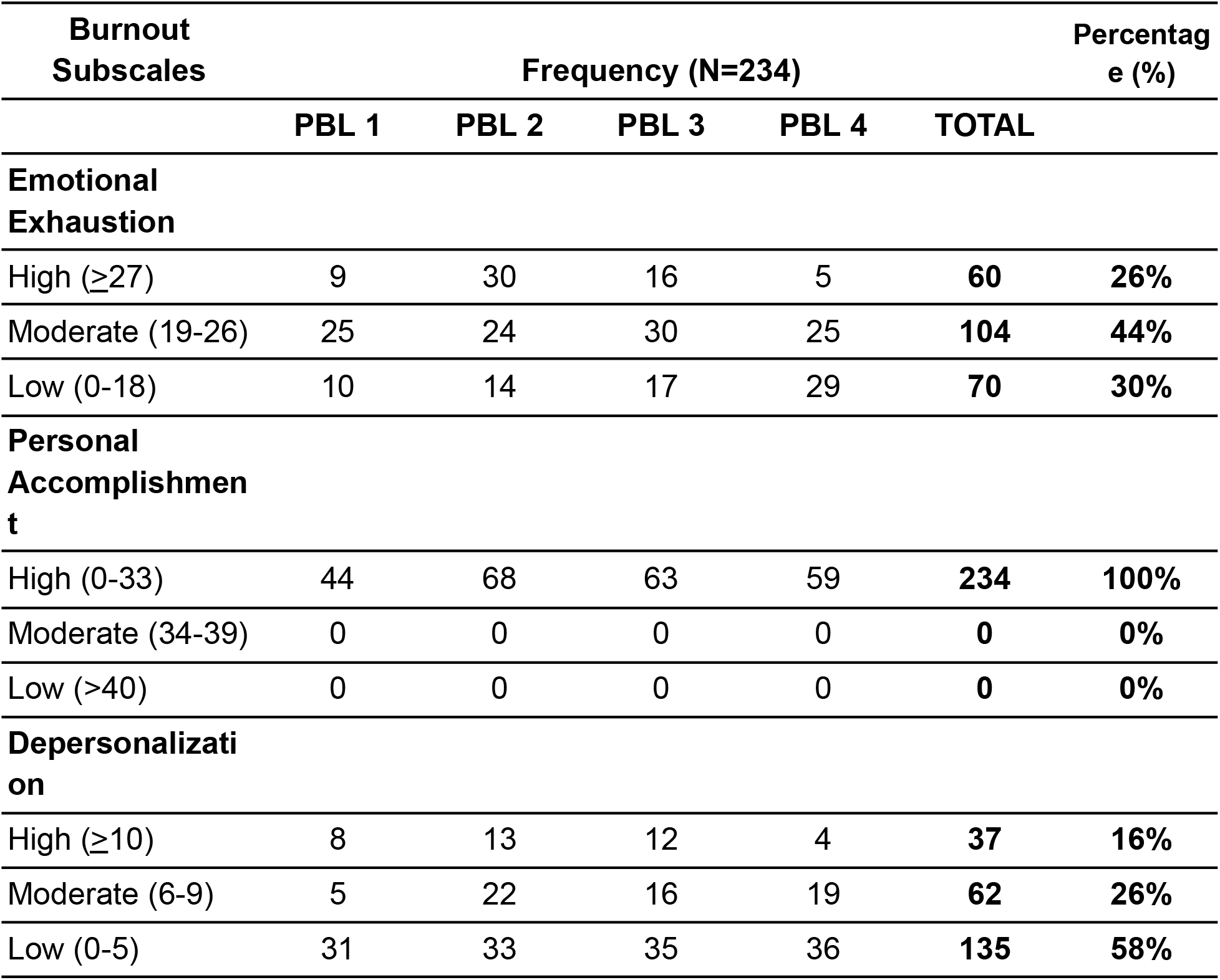
Frequency & Percentage Distribution Of The Level Of Burnout Of All Year Levels.

The overall incidence of burnout among selected medical students in all year levels is shown in Figure 1 and illustrates that PBL 2 has the highest incidence of burnout comprising 33 students (49%) while PBL 4 has the least burnout incidence with only 8 students (14%). It can also be inferred that the incidence of burnout decreases as the year level progresses, with the fourth year having the highest incidence of no burnout.

**Figure 1.**
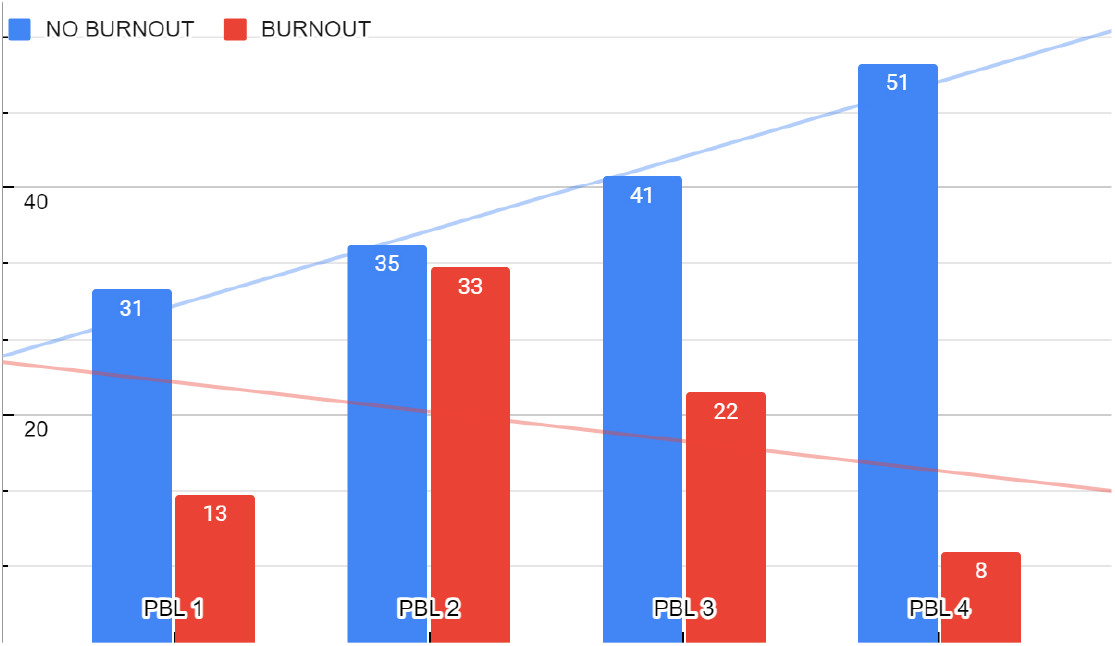
Medical Students Burnout Condition Across All Year Levels. (N=234)

**Figure 2.**
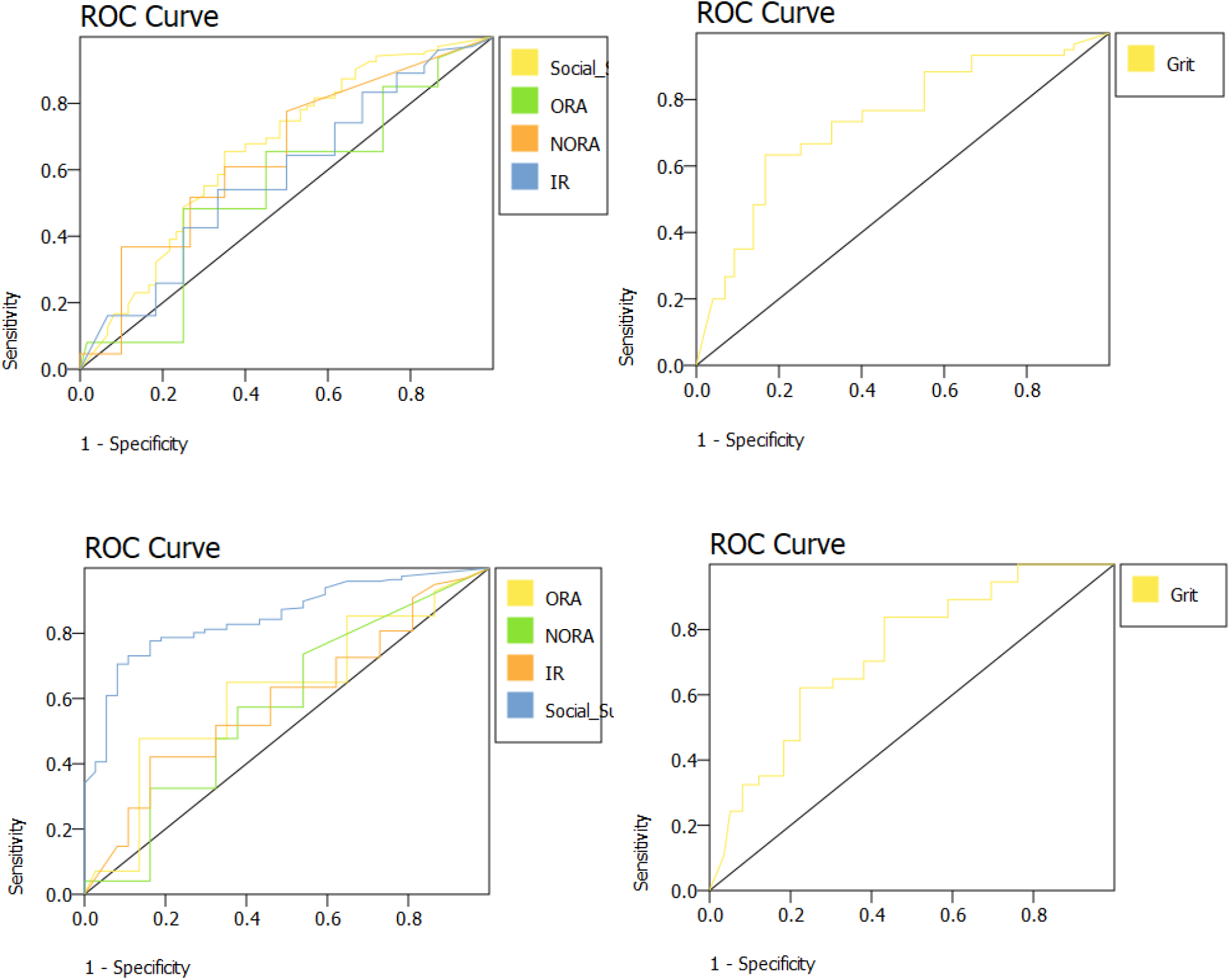
Receiver Operating Characteristic (ROC) Curve Analysis for grit, religiosity, and social support score as predictors of NO BURNOUT in the subscale of emotional exhaustion (above) and depersonalization (below).

In the analysis of specific protective factors, most medical students were shown to be moderately gritty (91%) while the rest are extremely gritty (8%) and not gritty (1%). Moreover, extreme grit was highest among the PBL 2 and PBL 4 students. This means that more gritty students are associated with lower levels of burnout. Most of the medical students are religious in the subscales of ORA and IR with 141 (60%) and 166 (71%) responses, respectively. These suggest that most respondents are involved or attend public religious activities. Additionally, the medical students also have a significant degree of personal religious commitment or motivation. However, most respondents have low scores in the NORA subscale (55%) suggesting no engagement in religious activities performed privately primarily attributed to the hectic schedules and time constraints of a medical student. Students with a high religiosity index had lower levels of psychological distress and burnout making it a protective factor. Majority of the medical students had high tolerance for ambiguity (98%). Tolerance for ambiguity is highest among PBL 2 students and lowest among PBL 1 students. Almost all medical students at all year levels tend to perceive ambiguous situations as desirable and have higher ambiguity tolerance resulting in improved emotional outcomes. In terms of the perceived social support of the respondents, most of the respondents have high social support from family (68%), friends (76%), and significant others (68%). From PBL 1 to 3, friends become increasingly important as a source of social support due to similar experiences and career choices as the respondents. For PBL 4 students, valuing family becomes more heightened because they are more exposed to real-life situations of patient-physician interactions and have developed family-orientedness. Perceived social support was found to significantly improve perceived sense of personal accomplishment hence reducing burnout level.

The ROC curves of grit, religiosity subdivided into ORA, NORA and IR, and social support were shown to be good predictors of burnout levels in the subscale of emotional exhaustion and depersonalization of selected medical students. Grit has good and acceptable diagnostic or prognostic accuracy and value to the presence or absence of the respective forms of burnouts while religiosity was considered moderately accurate predictors of burnout among medical students in all subscales. On the other hand, social support is a very good prognosticating marker of burnout with the highest AUC of 0.85 in the depersonalization subscale but it is not an acceptable prognosticator in the emotional exhaustion component of burnout. The specific protective resources that have significant utility in the management of burnout levels during the COVID-19 pandemic are grit, religiosity, and social support.

## CONCLUSION

Of the four protective resources, only grit, religiosity, and social support had a significant positive correlation to burnout while tolerance for ambiguity had no significant correlation to burnout. The three showed that they were good predictors of burnout levels - grit having good diagnostic accuracy and discrimination, while religiosity and social support being moderately accurate predictors of burnout.

## RECOMMENDATIONS

Recommendations for Future Studies:

1. Identify and explore relationships of other psychographic and social-demographic factors and burnout.
2. Conduct a prospective longitudinal study on medical students to clarify if the origin of the high levels of burnout syndrome are directly related to the progression of studies.
3. Evaluate and compare different protective resource assessment tools to determine the most appropriate tool for the future researchers in order to obtain the most precise results.
4. Conduct surveys to medical students in other Cebu medical schools with a different curriculums and determine any significant difference
5. Conduct surveys to students enrolled in a nonmedical field, and determine any significant difference
6. Formulate mental health wellness activities tailored to these specific protective resources to help medical students from developing negative coping strategies from burnout.

## Data Availability

All data produced in the present work are contained in the manuscript

